# What level of expertise is necessary to generate ACLS training test questions: pre-med students vs. artificial intelligence?

**DOI:** 10.64898/2026.06.11.26354470

**Authors:** Sunny LoGalbo, Mark Richman, Jeffrey Wang, Illan Saji, Aliyah Traore, Hannah Oliva, Evan Wu, Alessandro Drud, Diamia Foster, Sambhat Bhan dari, Raquel Lopez Delfillo, Amanda McCann, Jennifer Coard, Camille Matthew, Barry Smith

**Affiliations:** Northwell Health Long Island Jewish Medical Center, New Hyde Park, NY, United States of America

## Abstract

**Introduction:** In-hospital cardiac arrest carries high mortality despite standardized ACLS training. Educators face increasing time constraints in developing assessment tools for ACLS training. Two possible solutions to this problem are using pre-medical students or using artificial intelligence to generate test questions. This study compared the quality of pre-medical student-generated ACLS test questions vs. AI-generated ACLS test questions, testing the hypothesis that AI-generated questions are non-inferior to student-generated questions.

**Methods:** Ten pre-medical students created ACLS questions following predefined criteria, while an AI model (Northwell’s Artificial Intelligence Hub) generated comparable questions. A blinded ACLS-certified physician evaluated questions on the qualities of Alignment, Clarity, Cognitive Level, and Question Design using a standardized rubric (Likert scale:

1 = poor quality, 5 = excellent). Student’s T-test and Chi-square analysis were used to compare the quality of questions on different rubric domains within each arm (student vs. AI) and within one domain (eg, question Clarity) between arms. The Student’s T test was used when 2 comparator groups were compared (eg, Clarity of student-generated vs. AI-generated questions) within one arm. The ANOVA test was used when comparing more than 2 comparator groups (eg, Alignment vs. Clarity vs. Cognitive Level) within one arm. Statistical significance was set as a priority at p <0.05.

**Results:** Both student-generated and AI-generated questions were of high quality. AI-generated questions achieved the maximum score in the domains of Alignment, Clarity, and Question Design, but fell short of perfect scores in the domain of Cognitive Level (8 of 50 questions were less than 5). Student-generated questions achieved less-than-perfect scores in each domain. No significant difference was found in overall mean question scores between groups (students = 4.79, AI = 4.81; p = 0.9). However, AI-generated questions had significantly-greater Clarity (students = 4.8, AI = 5; p = .0461), while Alignment, Cognitive level, and Question Design showed no significant differences.

**Conclusion:** AI-generated questions demonstrated overall quality comparable to those generated by pre-medical students, supporting the potential role of AI as a scalable tool in ACLS educational assessment development. Further studies are warranted to evaluate additional AI platforms and determine optimal integration of AI in medical education assessment design.

## Introduction

In-hospital cardiac arrests occur approximately 290,000 times in adults each year in the United States. It is associated with a high mortality rate and requires rapid interventions to prevent, recognize, and reverse. Generally, outcomes are poor, with a survival-to-discharge rate of only about 25%.^1^ Improved outcomes are associated with early initiation of high quality Cardiopulmonary Resuscitation (CPR), and rapid defibrillation of certain arrhythmias.^2^ The American Heart Association (AHA) continuously develops standardized guidelines and evidence-based recommendations for managing cardiac arrest in both in-hospital and out-of-hospital settings, resulting in significant improvements in cardiac arrest outcomes.^3^ Advanced Cardiac Life Support (ACLS) training courses encompass these guidelines, and are required by essentially every level of healthcare provider (eg, medical doctor, physician assistant, nurse practitioner, emergency medicine technician - paramedic). ACLS trains providers in systematic approaches to treat cardiac arrest including, but not limited to, arrhythmia recognition, cardioversion, defibrillation, pharmacological management, and airway management.^4^

Clinician educators who organize and direct ACLS courses are increasingly-busy, with diminishing non-clinical time for assessing knowledge attained by learners.^5^ With less time available to dedicate to knowledge assessment attained during ACLS courses, it is important to derive a means of facilitating the creation of test questions that are valid, appropriately-challenging, and are not taken in whole or in part from existing ACLS tests available online. Two possible means for more-efficient creation of such test questions are pre-medical students (who either know ACLS or can learn about it online) and artificial intelligence (AI).

AI is currently being used in various other domains to generate medical examination questions and answers.^6^ However, there have been no studies examining the application of AI in the creation of ACLS training test questions and answers. Because AI has a tendency to “hallucinate^7^,” it may be unreliable in ACLS test question/answer generation compared with human-generated questions/answers. Therefore, it is essential to evaluate the quality of AI-generated ACLS test questions/answers in comparison to those created by humans. This study aims to investigate whether ACLS test questions/answers produced by AI are non-inferior to those produced by pre-medical students, as judged by an experienced, ACLS-trained physician. If AI is shown to generate inaccurate ACLS test questions or answers, while pre-medical students produce more-reliable content, academic physicians may feel more comfortable involving these students in ACLS question/answer development.

This study aims to compare the quality of AI-generated ACLS test questions/answers with those generated by pre-medical students. The hypothesis is that ACLS test questions are non-inferior to those generated by pre-medical students.

## Methods

Ten students created questions and answers (5 questions/answers each, for a total of 50 questions/answers). Subjects were recruited from Hofstra University courses held in collaboration with the Long Island Jewish Medical Center.

Subjects were invited to write 5 ACLS test questions/answers from one of the following:

- Bradycardia, including 1st-degree atrioventricular block (AVB), 2nd-degree Type I (Mobitz I) AVB, 2nd-degree Type II (Mobitz II), 3rd-degree AVB
- Narrow-complex tachycardia, including supraventricular tachycardia, atrial fibrillation, atrial flutter, multi-focal atrial tachycardia (MAT)
- Wide-complex tachycardia, including ventricular tachycardia and Torsade de Pointes
- Ventricular fibrillation
- Pulseless electrical activity and asystole

Subjects could choose which rhythm they wanted to write about, on a first-come, first-choice basis.

We gave subjects the following instructions describing characteristics/attributes of each set of 5 questions and answers:

- Subjects cannot use AI to write the questions and answers. They can use it to learn about the topic, but not to write the questions and answers.
- Pure knowledge vs. a case: Out of each subject’s 5 questions and answers, no more than 2 can be pure knowledge questions (eg, What are characteristics of 2nd degree Type I AV block?); the others must be narrative case questions.
- Multiple choice vs. true/false: Out of each subject’s 5 questions and answers, at least 3 must be multiple choice; there can be no more than 2 true/false questions. No questions can require a narrative response.
- True/false questions will have 2 response options (True vs. False); other questions will have 3-5 response options.
- Images vs. none: Out of each subject’s 5 questions and answers, at least 3 questions must contain an image of an EKG or rhythm strip.
- Intervention vs. diagnosis: Out of each subject’s 5 questions and answers, at least 3 must inquire about interventions (eg, what drug/dose to give, what Joules to shock at).
- If a subject chooses bradycardia: at least 2 questions and answers must be about 1st-degree AV block, 2 about Mobitz I AV block, 2 about Mobitz II AV block, and 2 about 3rd-degree AV block.
- If a subject chooses narrow-complex tachycardia: at least 2 questions and answers must be about SVT, 2 about atrial fibrillation, and 2 about atrial flutter.
- If a subject chooses wide-complex: at least 2 questions and answers must be about V tach and 2 questions and answers about Torsades de Pointes.
- If a subject chooses ventricular fibrillation, all questions and answers must be about ventricular fibrillation.
- If a subject chooses PEA/Asystole, all questions and answers must be about PEA/Asystole.

The AI source (Northwell’s Artificial Intelligence Hub) was prompted to create questions and answers that followed similar rules.

The ACLS-certified experienced physician (MR) judged these questions/answers according to a rubric, which contains guidelines pertaining to appropriateness of questions and accuracy of answers.

Key considerations:

1. Validity: The question/answer measures what it intends to measure
2. Reliability: The question/answer is straightforward
3. Conciseness: Unnecessary “fluff” language is removed to focus on the core concept

The rubric was based on best practices regarding writing questions and answers,^8^ using the following criteria that were assessed on a Likert scale (5 = excellent, 1 = poor).

Criteria domains:

1. Alignment: Directly measures a specific learning objective or standard
2. Clarity: Question/answer is concise, unambiguous, and easy to understand
3. Cognitive Level: Challenges higher-order thinking (application, analysis, evaluation)
4. Question Design: Distractors in multiple-choice questions are plausible, and there is only one answer that is indisputably-correct

The assessor was blinded as to whether the ACLS questions and answers were generated by a student or by the AI agent.

Study data were collected and managed using REDCap electronic data capture tools hosted at Northwell Health.^9,10^ REDCap (Research Electronic Data Capture) is a secure, web-based software platform designed to support data capture for research studies, providing 1) an intuitive interface for validated data capture; 2) audit trails for tracking data manipulation and export procedures; 3) automated export procedures for seamless data downloads to common statistical packages; and 4) procedures for data integration and interoperability with external sources.

### Statistics

We compared differences in mean score and percent of scores <5 between quality domains (eg, Alignment vs. Clarity vs. Cognitive Level) within one arm (eg, within the student arm, and, separately, within the AI arm). We then compared each separate quality domain between arms (eg, Alignment of student-generated vs. AI-generated questions/answers; Clarity of student-generated vs. AI-generated questions/answers). The Student’s T test was used when 2 comparator groups were compared (eg, Clarity of student-generated vs. AI-generated questions/answers) within one arm. The ANOVA test was used when comparing more than 2 comparator groups (eg, Alignment vs. Clarity vs. Cognitive Level) within one arm.^11,12^ Statistical significance was set as a priority at p <0.05.

**This project was reviewed and deemed not to meet the definition of research by the Northwell Health Institutional Review Board’s (IRB’s) Human Research Protection Program (HSRD24-0133), indicating that formal IRB approval was not required for this study. All observations were collected in compliance with institutional guidelines for patient privacy and data security**

## Results

The quality of student questions/answers varied by domain (eg, Alignment, Clarity). As shown in Table 1, 2.0% of questions/answers had a score in the Alignment domain <5, 8.0% had a Clarity score <5, 16.0% had a Cognitive Level score of <5, and 6.0% had a Design score <5 (*p <0*.*05)*. Questions/answers tended to be well-aligned with the learning objective (mean = 4.94), but the Cognitive Level of several questions/answers was less-than-rigorous (mean = 4.54). No AI-generated questions/answers exhibited “hallucinations.”

**Table 1.** Student ACLS Question Quality.

| **Value** | **Alignment<sup>a</sup>** | **Clarity<sup>b</sup>** | **Cognitive Level<sup>c</sup>** | **Question Design<sup>d</sup>** | **P value** |
| --- | --- | --- | --- | --- | --- |
| Mean | 4.94 | 4.8 | 4.54 | 4.86 | 0.048 |
| % with score <5 | 2.0% | 8.0% | 16.0% | 6.0% |  |
| <sup>a</sup>Directly measures a specific learning objective or standard |  |  |  |  |  |
| <sup>b</sup>Question is concise, unambiguous, and easy to understand |  |  |  |  |  |
| <sup>c</sup>Challenges higher-order thinking (application, analysis, evaluation) |  |  |  |  |  |
| <sup>d</sup>Distractors in multiple-choice questions are plausible, and the answer is indisputably correct |  |  |  |  |  |

**Table 2.**
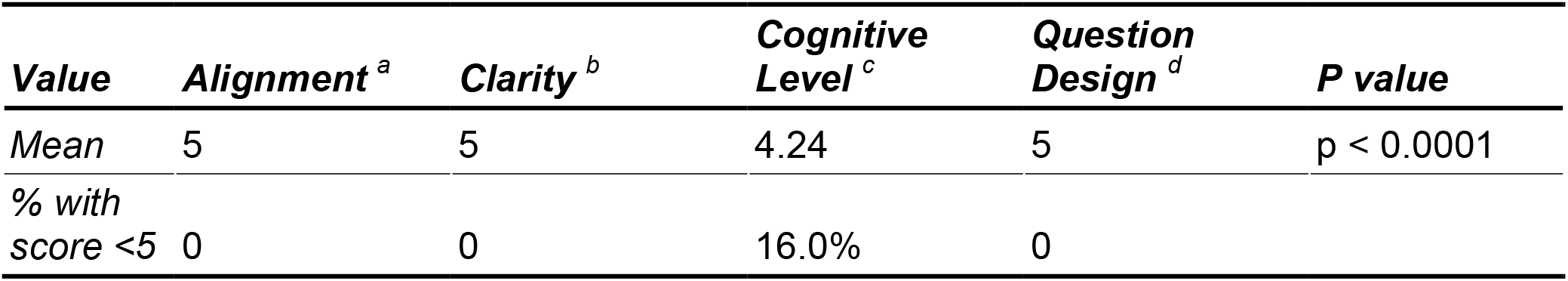
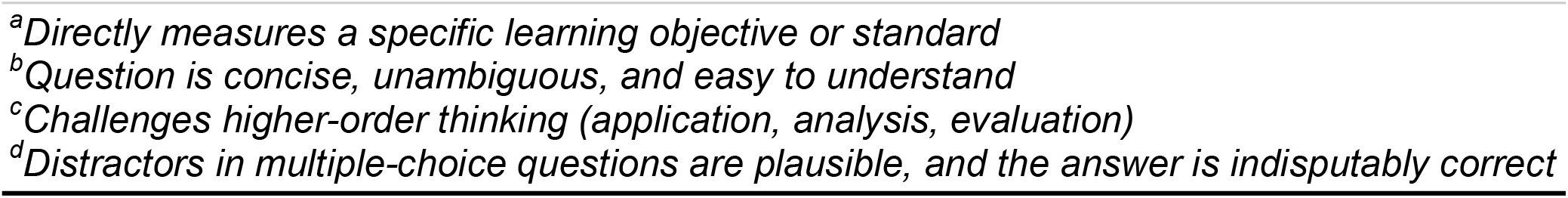
AI ACLS Question Quality.

| **Value** | **Alignment<sup>a</sup>** | **Clarity<sup>b</sup>** | **Cognitive Level<sup>c</sup>** | **Question Design<sup>d</sup>** | **P value** |
| --- | --- | --- | --- | --- | --- |
| Mean | 5 | 5 | 4.24 | 5 | $p < 0.0001$ |
| % with score <5 | 0 | 0 | 16.0% | 0 |  |

<sup>a</sup>Directly measures a specific learning objective or standard
<sup>b</sup>Question is concise, unambiguous, and easy to understand
<sup>c</sup>Challenges higher-order thinking (application, analysis, evaluation)
<sup>d</sup>Distractors in multiple-choice questions are plausible, and the answer is indisputably correct

In contrast, AI-generated ACLS questions/answers were excellent in Alignment, Clarity, and Design (100% of questions/answers were rated a 5 in these domains), but suffered disproportionately in Cognitive Level (mean score 4.24, with 16.0% having a Cognitive Level score <5). Several were quite simple. For example, a question would show an EKG and ask “Which of the following BEST describes this rhythm?**”** No questions had scores <5 in any other domain of evaluation (p <0.0001).

Comparison of the means in each domain between the student-created and AI-created questions/answers revealed a statistically-significant difference only in the Clarity domain (p = 0.046). (**Table 3, Figure 1**).

**Table 3.** Comparison of the Means for Student-Created and AI-Created Questions by Question Domain.

| **Value** | **Alignment<sup>a</sup>** | **Clarity<sup>b</sup>** | **Cognitive Level<sup>c</sup>** | **Question Design<sup>d</sup>** | **P value** |
| --- | --- | --- | --- | --- | --- |
| Student Mean | 4.94 | 4.8 | 4.54 | 4.86 | 0.048 |
| AI Mean | 5 | 5 | 4.24 | 5 | <0.0001 |
| P-Value | 0.3149 | 0.0461* | 0.1901 | 0.1251 |  |

<sup>a</sup>Directly measures a specific learning objective or standard
<sup>b</sup>Question is concise, unambiguous, and easy to understand
<sup>c</sup>Challenges higher-order thinking (application, analysis, evaluation)
<sup>d</sup>Distractors in multiple-choice questions are plausible, and the answer is indisputably correct
\*Statistically Significant ( $p < 0.05$ )

**Figure 1.**
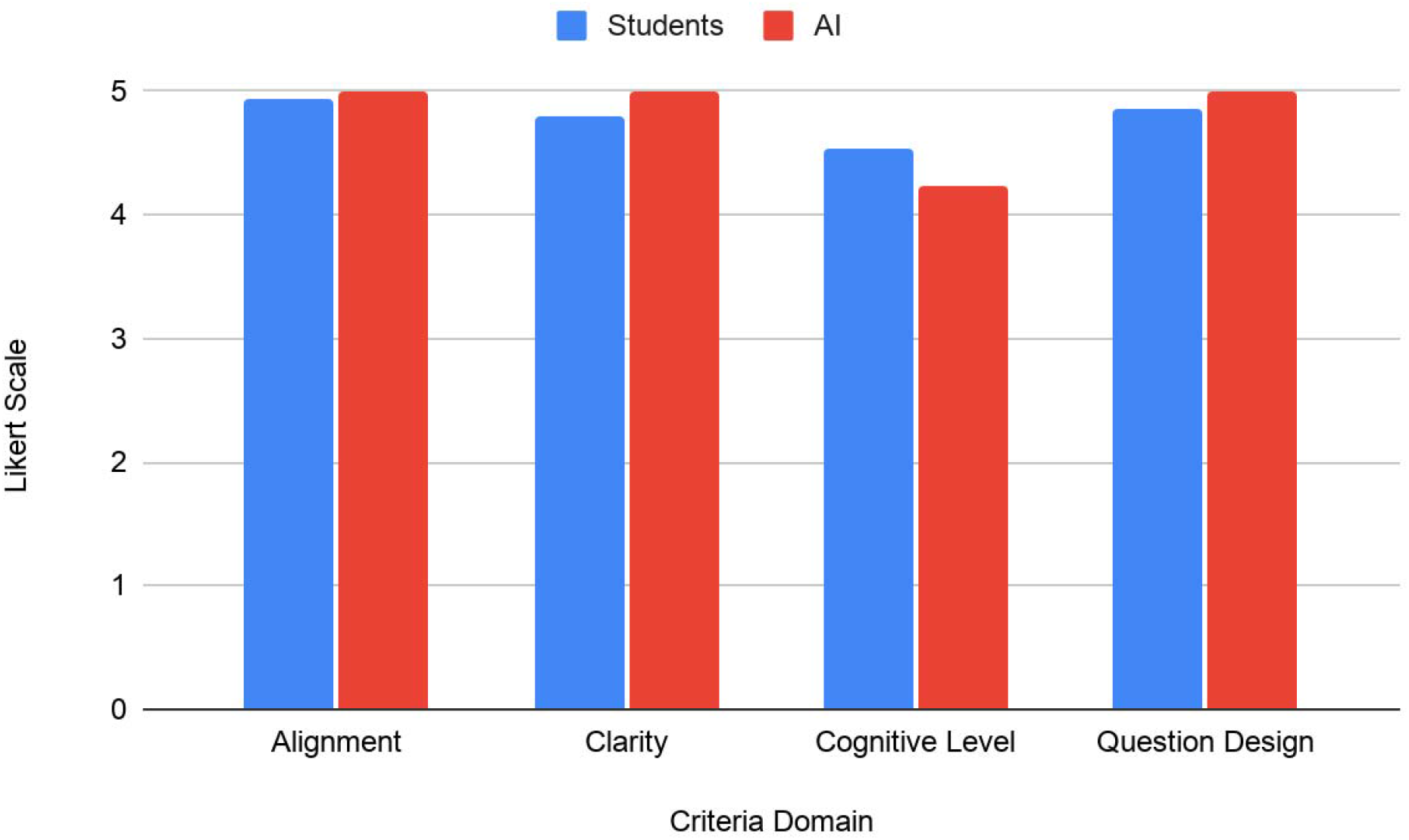
Student’s vs. AI’s ACLS Question Quality

## Discussion

Comparison between student- and AI-generated questions/answers did not differ in the domains of Alignment, Cognitive Level, and Question Design. However, in the Clarity domain, the AI-generated questions were significantly-better than student-generated questions (p <0.05).

AI-generated questions/answers were rated to have perfect scores in the Alignment, Clarity, or Question Design domains. The sole domain in which AI-generated questions did not score a maximum of 5 was Cognitive Level. Some questions were too simple and did not require enough higher-order thinking or knowledge.

This study demonstrates AI is effective in generating high-quality ACLS certification examination questions/answers, with the caveat that the questions/answers must be screened for difficulty. However, use of AI for such purpose would still decrease the clinician educator’s overall workload, which could help to address the issue of overburdening. Pre-medical student questions/answers were also of high quality; however, having students create questions/answers would create an additional workload burden for such students.

### Limitations

This was a single-site study with pre-medical students recruited from only one university. In addition, AI models are rapidly improving, and the quality of their questions/answers in the domain of Cognitive Level might achieve the maximum rating of 5 in the near term. Finally, we only tested the Google Gemini 2.5 Flash model, though there are multiple other publicly-available models. Future researchers may wish to compare question/answer quality between models.

## Data Availability

All data produced in the present study are available upon reasonable request to the authors

